# In-house modification and improvement of the CDC real-time PCR diagnostic assay for SARS-CoV-2 detection

**DOI:** 10.1101/2020.07.10.20150771

**Authors:** Srirupa Das, Candice Dowell-Martino, Lisa Arrigo, Paul N. Fiedler, Sandra Lobo

**Affiliations:** Department of Pathology Research, Nuvance Health, Danbury, CT 06810, USA; Department of Pancreatic Research, Nuvance Health, Danbury, CT 06810, USA; Department of Pathology, Nuvance Health, Danbury, CT 06810, USA; Rudy L. Ruggles Biomedical Research Institute, Nuvance Health, Danbury, CT 06810, USA

**Keywords:** SARS-CoV-2, COVID-19, Real-time PCR, Centers for Disease Control and Prevention (CDC), Diagnostic assay

## Abstract

The world is currently facing an unprecedented pandemic caused by the novel coronavirus SARS-CoV-2 (COVID-19) which was first reported in late 2019 by China to the World Health Organization (WHO). The containment strategy for COVID-19, which has non-specific flu-like symptoms and where upwards of 80% of the affected has either mild or no symptoms, is critically centered upon diagnostic testing, tracking and isolation. Thus, the development of specific and sensitive diagnostic tests for COVID-19 is key towards the first successful step of disease management. Public health organizations like the WHO and the US-based Centers for Disease Control and Prevention (CDC) have developed real-time PCR (RT-PCR) based diagnostic tests to aid in the detection of acute infection. In this study we sought to modify the CDC RT-PCR diagnostic assay protocol to increase its sensitivity and to make the assay directly portable to health care providers in a community-based hospital setting. A number of modifications to the original protocol were tested. Increasing the RT-PCR annealing temperature by 7°C to 62°C was associated with the most significant improvement in sensitivity, wherein the cycle-threshold (Ct) value for the N2 assay was reduced by ∼3 units, in effect both reducing the overall number of inconclusive results and yielding N1/N2 assays to have similar Ct values. The limit of detection of the modified assay was also improved (0.86 RNA copies/µl for both nCoV 2019_N1/N2 assays) compared to the CDC RT-PCR diagnostic assay (1 and 3.16 RNA copies/µl for nCoV 2019_N1 and N2 assay, respectively). Using this modification, there was no significant effect on SARS-CoV-2 detection rate when viral RNA extraction was performed either manually or through an automated extraction method. We believe this modified protocol allows for more sensitive detection of the virus which in turn will be useful for pandemic management.

## Introduction

The initial outbreak of a severe respiratory disease in China caused by a novel coronavirus of unknown origin was first reported to the World Health Organization (WHO) on December 31, 2019 [1, 2]. In this first half year since its reporting, and at the time of this writing, this disease has spread to 188 countries with more than 12.07 million documented cases and more than 550,000 confirmed deaths worldwide [3]. The US reported its first confirmed case of a person-to-person transmission of this novel disease on January 30^th^, 2020 [2] and since then the US has been the worst affected country in the world with more than 3 million confirmed cases and >130,000 deaths [3]. These numbers are most likely vastly underreported than the actual number of infected and virus-attributable deaths and will continue to grow for the foreseeable future. There are seven known species of coronaviruses that infect humans: 229E, NL63, OC43, HKU1, MERS-CoV, SARS-CoV-1 and now the novel coronavirus that causes COVID-19, named SARS-CoV-2 [4]. The first four coronavirus strains cause mild symptoms as compared to the more recent strains like MERS-CoV, the etiologic agent of the Middle East Respiratory Syndrome outbreak of 2012; SARS-CoV-1 responsible for the Severe Acute Respiratory Syndrome epidemic of 2002-2003 and SARS-CoV-2, responsible for the deadly COVID-19 pandemic [4].

The blinding speed with which the COVID-19 has spread across the world, causing thousands of deaths within a short span of time and impacting millions, has put a lot of pressure on federal governments and public health organizations in terms of disease management and preparedness to handle a pandemic of this magnitude. The US Centers for Disease Control and Prevention (CDC) developed a laboratory test kit for a real-time PCR (Polymerase Chain Reaction)-based diagnostic test, the “Centers for Disease Control and Prevention (CDC) 2019 Novel Coronavirus (2019-nCoV) Real-Time Reverse Transcriptase (RT)–PCR Diagnostic Panel.” [5]. Since then, many laboratories and organizations have either employed this exact test or developed their own modified forms to enhance testing capability. Given that diagnostic testing is the central tenet to understanding and minimizing disease spread, the US Food and Drug Administration (FDA) relaxed their own existing rules and issued emergency use authorizations (EUAs) to help cope with the unprecedented demand for diagnostic testing. At the time of this writing, the FDA has issued EUAs to 141 *in vitro* diagnostic tests for COVID-19 including several antibody-based tests [6].

Evidence suggests that 70-80% of COVID-19 patients are asymptomatic or have very mild symptoms [7]. All affecteds are contagious and thus capable of spreading the virus. The critical initial steps in disease management begin with testing, tracking and isolation [8, 9]. Accurate and specific diagnostic testing for SARS-CoV-2 is the pillar upon which this disease management pipeline is based. During the first peak of the US COVID-19 pandemic, there were shortages of testing and large commercial laboratories reported multi-day back logs for reporting results, which rendered patient management ineffective and resulted in multiple inefficiencies of resources but most importantly, possibly contributed to increased disease spread and death. Due to these delays in COVID-19 testing, major community health network like Nuvance Health, a group of seven hospitals in the states of Connecticut and New York, located in the US epicenter of the COVID-19 outbreak needed to develop their own diagnostic test that can be easily offered to hundreds of patients that come through the system every day and to the frontline healthcare workers.

Testing for COVID-19 is not feasible by viral culture as the virus is respiratory in nature, highly contagious and requires biosafety level 3 facilities that are not available in most health-care institutions. Serology-based testing although safer to perform, can give a higher false negative as the test is dependent on the host mounting an immune response post-infection that can take some time, making this test unsuitable for tracking acute infections. PCR is considered to be the “gold-standard” in molecular diagnostic testing and since its invention in 1983 has been used in the development of several diagnostic tests for infectious diseases [10]. Availability of the complete SARS-CoV-2 genome sequence early in the pandemic [11] helped to facilitate the development of the PCR-based diagnostic tests by the WHO [12] and the CDC [5]. We report on our modification of the CDC-based assay which was refined with the dual aims of developing a rapid, high sensitivity test that could be quickly ported into clinical use in a community-based, hospital network.

## Material and Methods

### Primers and probes

The primers and probes used in this study were purchased from Integrated DNA Technologies (IDT; Cat No. 10006713), IA (Table 1). These primers and probes were based upon the sequences provided by the CDC EUA protocol for COVID-19 diagnostic testing [5]. The N1 and N2 assays were designed based on the Nucleocapsid gene (N) sequence of the SARS-CoV-2 virus (Gene Bank Accession No. NC_045512.2). The human RNase P (Hs_RP) assay was included as an extraction control.

**Table 1:**
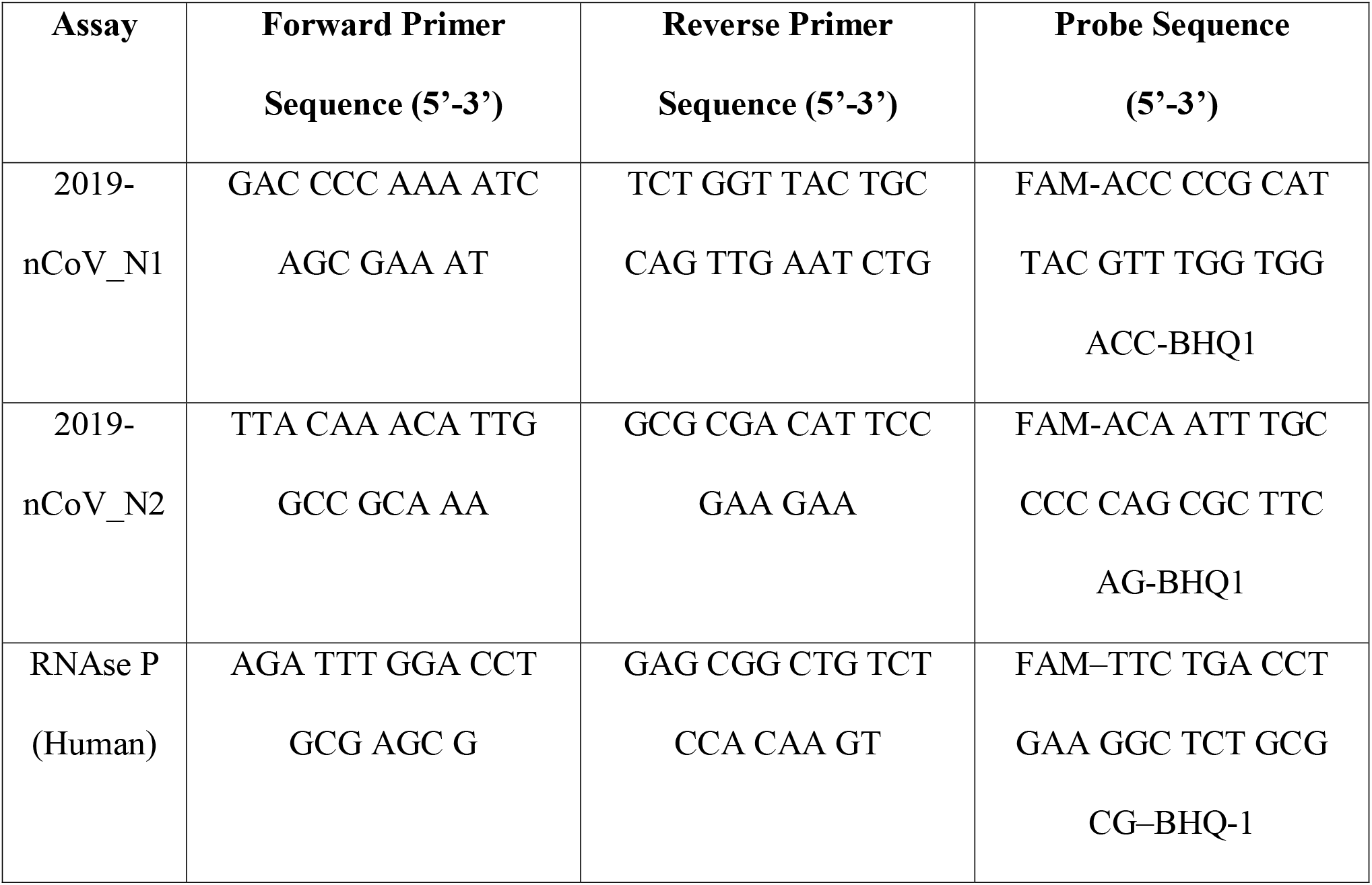
TaqMan primer and probe sequences.

### Control reagents

Positive and negative control reagents used for development of the assay were purchased from IDT, IA. The positive plasmid control reagents used were: 2019-nCoV_N_Positive Control (Cat No. 10006625) and Hs_RPP30 Positive Control (Cat N0. 10006626). The negative control plasmids were: MERS-CoV Control (Cat No. 10006623) and SARS-CoV Control (Cat No. 10006624). Quantitative Synthetic SARS-CoV-2 RNA: ORF, E, N (ATCC^®^ VR-3276SD™) was used for all limit of detection (LOD) experiments. All dilutions were prepared using a DNA suspension buffer (Teknova, CA).

### Cell culture

The adenocarcinoma human alveolar basal epithelial cell line A549 was purchased from ATCC, Manassas, VA and cultured in 1640-RPMI medium supplemented with 10% heat inactivated fetal bovine serum, 2mM L-glutamine and 1% penicillin-streptomycin (Thermo-Fisher Scientific, Waltham, MA) for complete media. Cells were grown in an air-humidified incubator at 37°C with 5% CO_2_. Culture media was replaced every two days and cells were passaged at 80% confluence. For various experiments, the confluent monolayer was trypsinized using 0.25% trypsin/EDTA (Thermo-Fisher Scientific, Waltham, MA), collected and then centrifuged at 500g for 7 minutes. The cell pellet was washed twice with sterile 1X phosphate-buffered saline (Thermo-Fisher Scientific, Waltham, MA) for downstream applications. A549 cells were either pelleted for total RNA extraction or stored in viral transport medium (VTM) for preparing samples for LOD experiments. To define cell concentrations, cells were first mixed (1:1) with Trypan blue reagent (Thermo-Fisher Scientific, Waltham, MA) and then these suspended cells subjected to automated number and viability assessment using the Countess Automated Cell Counter (Life Technologies, Carlsbad, CA).

### Total human RNA extraction

Total human RNA was purified from 1 million A549 cells using RNeasy Purification Kit (Qiagen, Hilden, Germany) according to the manufacturer’s specifications. The yield and quality of the RNA was assessed using Nanodrop Spectrophotometer (Thermo-Fisher Scientific, Waltham, MA, USA).

### Viral Transport Media preparation

VTM was prepared in accordance to the CDC recommended protocol [13].

### Sample preparation and viral RNA extraction for LOD determination

LOD studies of SARS-CoV-2 were performed using a quantitative synthetic SARS-CoV-2 RNA (ATCC, Cat No. VR-3276SD) that was spiked into a diluent consisting of a suspension of human A549 cells (500 cells) and 140µl of VTM, to mimic a positive clinical specimen. The synthetic RNA stock was 3-fold serially diluted with DNA suspension buffer containing 2ng/µl of carrier RNA (Qiagen, Hilden, Germany), to form a dilution series such that when 5µl of each dilution was spiked, a range of 15,000 copies (equivalent of 103.4 copies/µl, as per the CDC EUA) to 50 copies (equivalent to 0.34 copies/µl according to the CDC) could be approximated [5]. All dilutions were performed on ice and in a sterile cabinet. Samples were extracted using the QIAamp Viral RNA Mini Kit (Qiagen, Hilden, Germany), either using the automated QIAcube (Qiagen, Hilden, Germany) or manually, according to the vendor’s instructions. The lysis step prior to RNA extraction for all samples was performed manually by the addition of 560µl of Buffer AVL into each micro-centrifuge tube, followed by the addition of 140µl of the clinical matrix (A549 cells in VTM) and lastly, 5µl of the appropriate synthetic RNA spike amount. All samples were incubated for 10 minutes at room temperature. The remainder of the steps in the RNA extraction protocol was either performed manually or automated using the QIAcube. All extracted samples were eluted in a final volume of 100µl and immediately processed for downstream PCR.

### Quantitative real-time PCR (qPCR) protocol

PCR was performed using a QuantStudio 7 Flex Real Time PCR System (Life Technologies, Carlsbad, CA), following the manufacturer’s instructions. QPCR was performed in a total final reaction volume of 20µl with 5µl of template DNA or RNA, 500nM of primer/probe mix and 1X TaqPath 1-step RT-qPCR master mix, CG (Life-Technologies, Carlsbad, California). Nuclease-free water was added to complete the remaining volume. The CDC recommended PCR parameters were first used: 25°C - 2 min; 50°C - 15 min; 95°C - 2min; 45 cycles of: 95°C - 3 sec, 62°C - 30 sec [5]. The annealing temperature and primer/probe concentration were varied to optimize the PCR reaction, as described in the Results section. A cycle threshold (Ct) value of less than 40 (as per CDC recommendations) was considered to be positive for detection [5]. All PCR reactions were performed in triplicate and repeated three times.

## Results

### Analytical sensitivity and specificity testing of SARS-CoV-2 and human-specific gene assays with DNA controls

As a first step towards assay development, we began by testing the analytical sensitivity of each TaqMan assay for the SARS-CoV-2 viral N gene (N1 and N2) and the human RNase P gene (Hs_RP). This was determined using serial two-fold serial dilutions of the positive plasmid control DNAs (Table 2). CDC recommended PCR parameters were used with the following DNA concentrations: 100, 50, 25, 12.5, 6.25, 3.125 and 1.562 copies. The assay had analytical LOD of 6.25 DNA copies of SARS-CoV-2 for the N1 assay and 50 DNA copies for the N2 assay (Table 2). When lower copy numbers of DNA were tested, qPCR detection failed due to low statistical probability of incorporation of these low copy number DNA targets into the PCR reaction. According to the CDC interpretation guide for patient samples, Ct values of greater than 40 are considered as negative [5]. For the human RNase P gene, the assay had a sensitivity of 3.125 DNA copies (Table 2).

**Table 2:**
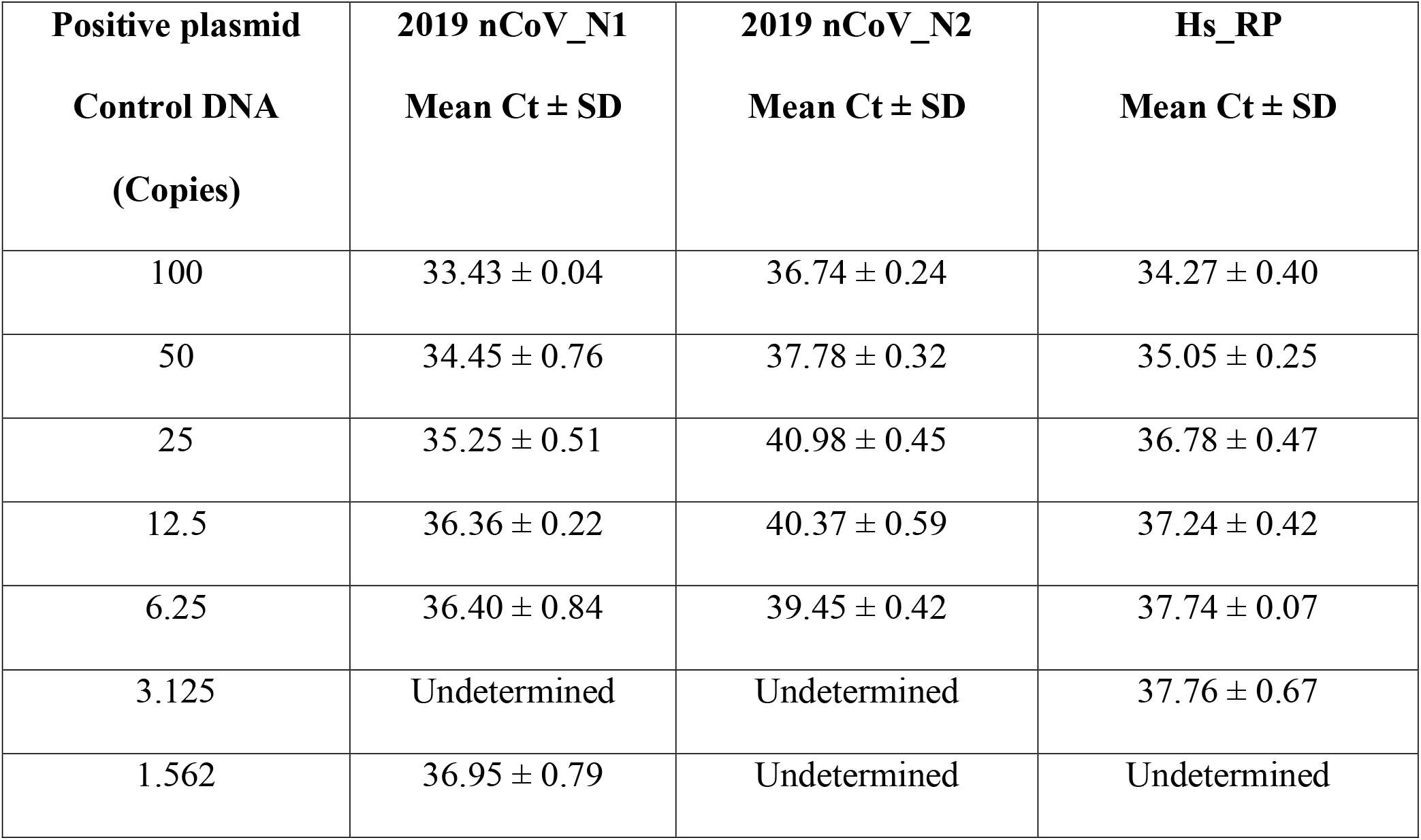
Analytical sensitivity using positive plasmid control DNA.

| **Positive plasmid**<br>**Control DNA**<br>**(Copies)** | **2019 nCoV_N1**<br>**Mean Ct ± SD** | **2019 nCoV_N2**<br>**Mean Ct ± SD** | **Hs_RP**<br>**Mean Ct ± SD** |
| --- | --- | --- | --- |
| 100 | 33.43 ± 0.04 | 36.74 ± 0.24 | 34.27 ± 0.40 |
| 50 | 34.45 ± 0.76 | 37.78 ± 0.32 | 35.05 ± 0.25 |
| 25 | 35.25 ± 0.51 | 40.98 ± 0.45 | 36.78 ± 0.47 |
| 12.5 | 36.36 ± 0.22 | 40.37 ± 0.59 | 37.24 ± 0.42 |
| 6.25 | 36.40 ± 0.84 | 39.45 ± 0.42 | 37.74 ± 0.07 |
| 3.125 | Undetermined | Undetermined | 37.76 ± 0.67 |
| 1.562 | 36.95 ± 0.79 | Undetermined | Undetermined |

Analytical specificity testing of the TaqMan assays for the SARS-CoV-2 viral N gene (N1 and N2) and the human RNase P gene (Hs_RP) was performed by qPCR with negative plasmid control DNAs (100 genome copies of MERS-CoV and SARS-CoV) and with 1ng of total DNA from 27 clinically relevant microbial species likely to be encountered in a human patient sample, as listed in Supplemental Table 1. Total genomic DNA from these pathogens was extracted by QIAamp DNA mini kit (Qiagen Hilden, Germany), according to the vendor’s instructions and quantitated by Nanodrop Spectrophotometer (Thermo Fisher Scientific, Waltham, MA, USA). DNA from 2019-nCoV_N_Positive Control (IDT#10006625) and Hs_RPP30 Positive Control (IDT#10006626) served as positive controls in the assay. None of the TaqMan assays demonstrated cross-reactivity with any of the control pathogenic agents tested (data not shown).

### Optimization of PCR conditions

To ensure robustness of the PCR assay, a number of modifiable parameters were selected for testing using positive plasmid DNA controls (either 50 or 100 copies per PCR reaction). These parameters most notably included the concentration of the primer/probe mixes and qPCR annealing temperatures. A positive effect was considered achieved if Ct values were decreased. Alteration of the primer/probe concentration from the CDC recommended concentration of 500nM of primer and 125nM of probe, while holding the other qPCR assay parameters as recommended by the CDC protocol [5], did not have any significant effect on the Ct values for any of the nCoV 2019_N1, nCoV 2019_N2 and Hs_RP assays (Supplemental Table 2). We then interrogated the effect of changing annealing temperatures. Including the recommended annealing temperature of 55°C, and based on our calculations of melting temperatures through sequence analysis of the primers being used, we tested across a broad range of temperatures: 53°C, 55°C, 57°C, 60°C and 62°C. A significant difference in Ct values, especially for the N2 assay (Table 3) was identified. The Ct values were the lowest at our maximum temperature of 62°C for the N2 assay and gradually increased with lower annealing temperatures (Table 3). The Ct values for the N2 assay at 62°C annealing temperature was lower by ∼3 units, as compared to the CDC recommended annealing temperature of 55°C. However, for the N1 and the human RNase P assays there were no significant difference in the Ct values across the entire range of the annealing temperatures (Table 3). We therefore chose 62°C as our assay annealing temperature.

**Table 3:**
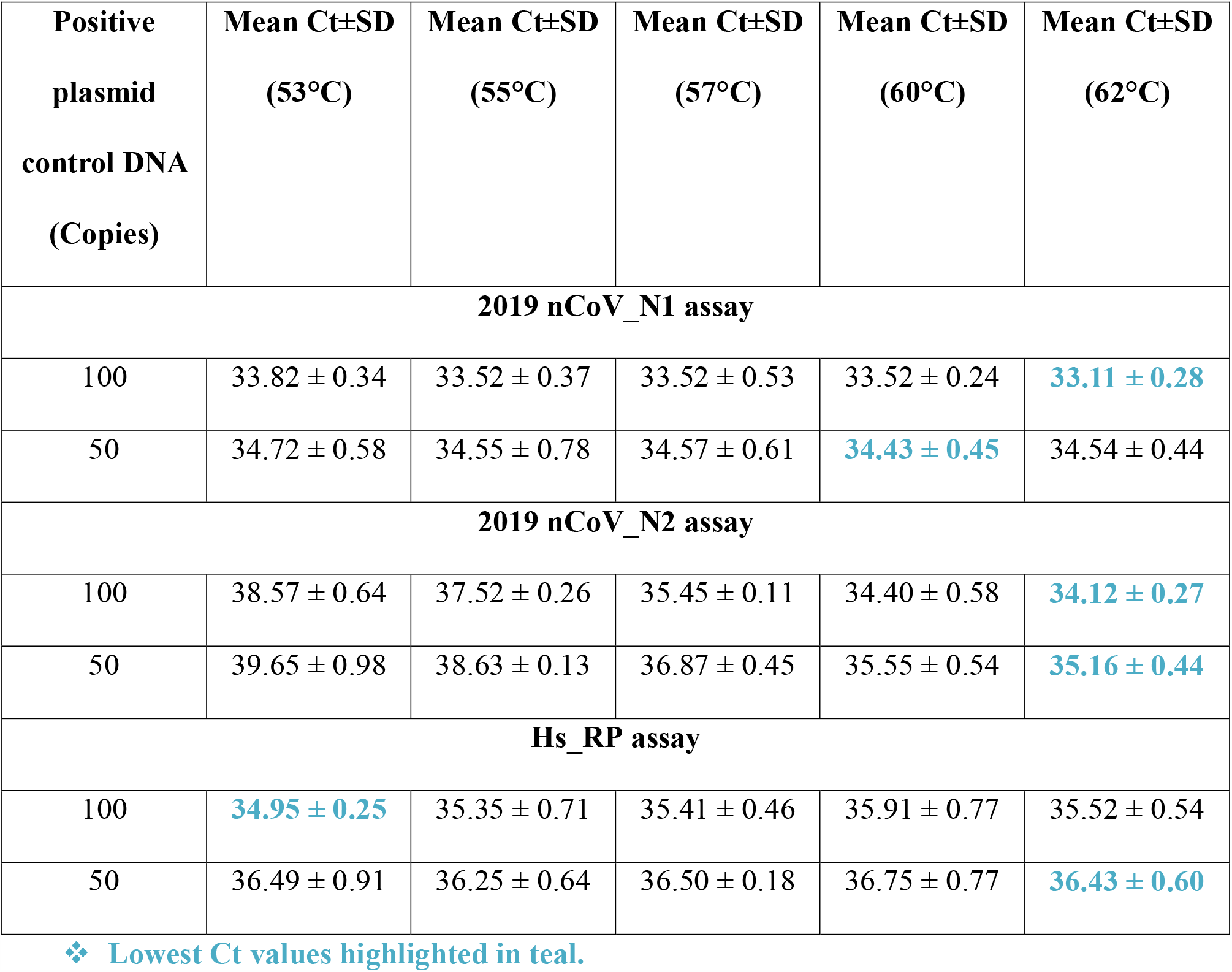
Optimization of PCR assay annealing temperature.

### Linearity range and LOD determination

To determine the linear dynamic range of the qPCR assay, 10-fold serial dilutions of positive control DNAs were used to create six concentration levels (range: 10^6^ - 10 copies/reaction) and then tested with both lab-optimized and CDC-recommended PCR parameters. The lab-optimized PCR parameters yielded a linear dynamic range across the entire tested dilution spectrum and reliably detected even 10 copies of both the N1 and N2 targets (Figure 1). In contrast, although the CDC recommended temperature conditions yielded a linear range across the spectrum, the Ct values for the N2 assay was more than 40 for 100 copies and other lower concentrations (Supplemental Figure 1).

**Figure 1.**
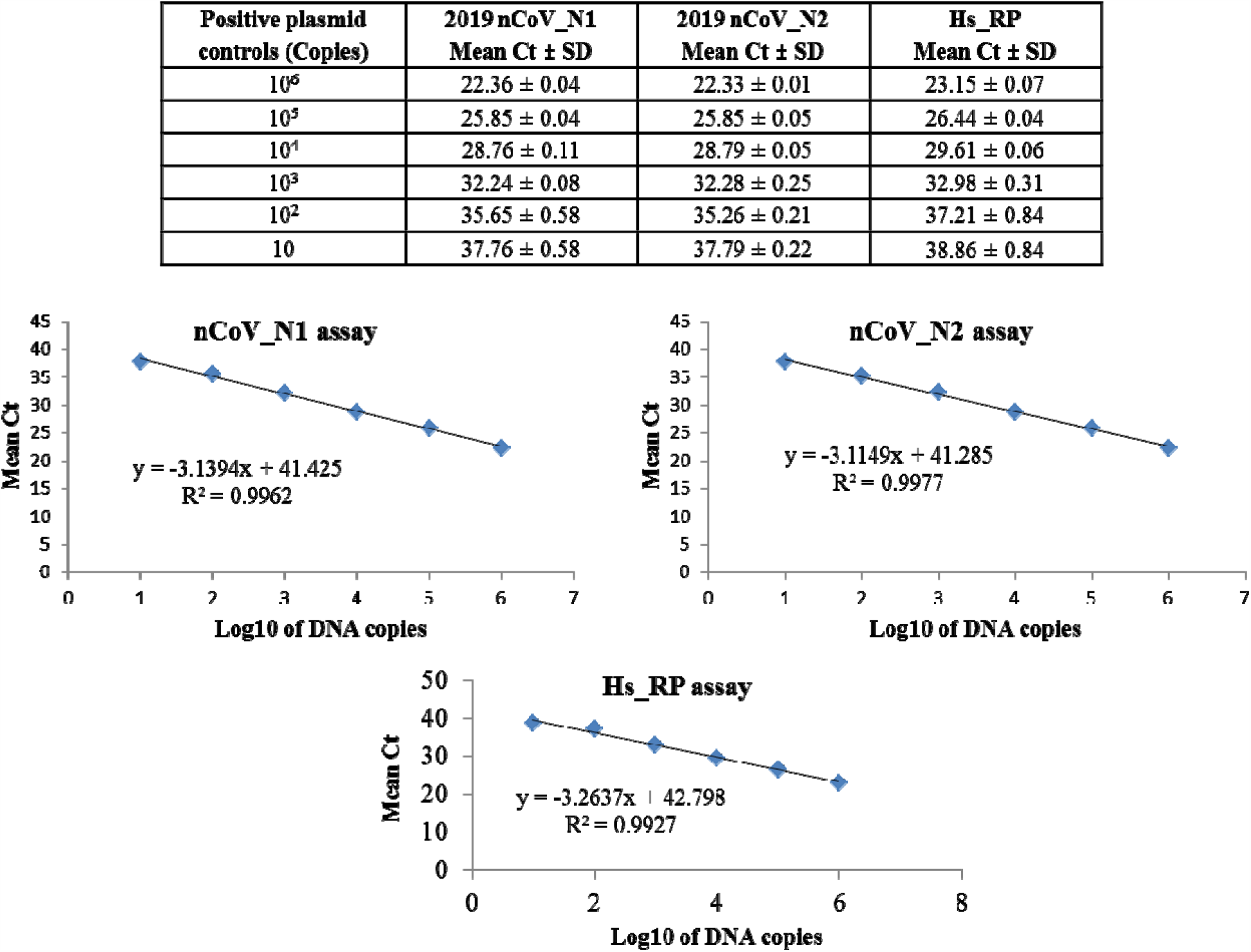
Dynamic linear real-time PCR range with lab-optimized PCR conditions. The linearity range of 2019 nCoV_N1, 2019 nCoV_N2 and Hs_RP Taqman assays wa determined with lab-optimized PCR parameters and various concentrations of positive plasmid control DNA by real-time PCR.

The LOD was determined twice for the COVID-19 diagnostic assay: with and without the viral RNA extraction step. LOD is the lowest concentration that is detected ≥95% of the times they are tested. For LOD determination without the RNA extraction step, quantitative synthetic SARS-CoV-2 RNA (ATCC#VR-3276SD) was serially diluted two-fold in DNA suspension buffer (with 2ng/µl carrier RNA) and then spiked into 2.5µg of total RNA (per PCR reaction) from human A549 cells, to mimic a clinical sample. The concentration range used for LOD determination was: 100, 50, 25, 12.5, 6.25, 3.125 and 1.562 copies of SARS-CoV-2 synthetic RNA. PCR was performed comparing CDC and lab optimized temperature parameters. As shown in Table 4, theoretically the LOD was 1.562 copies of the SARS-CoV-2 synthetic RNA for both N1 and N2 assays under the lab optimized conditions. Given the low Ct values for the 1.562 RNA copies for the N1 assay-37.18 and the N2 assay-37.13 (Table 4) we can in all likelihood detect even lower copy numbers before hitting the threshold Ct value of 40, as per CDC guidelines [5]. By comparison, under the CDC recommended conditions, the N2 assay had a LOD of 3.125 copies of synthetic RNA (Table 4). Two different operators repeated the experiment to determine operator-to-operator variability which was found to be <5% for the N1 assay and <2% for the N2 assay (data not shown).

**Table 4:**
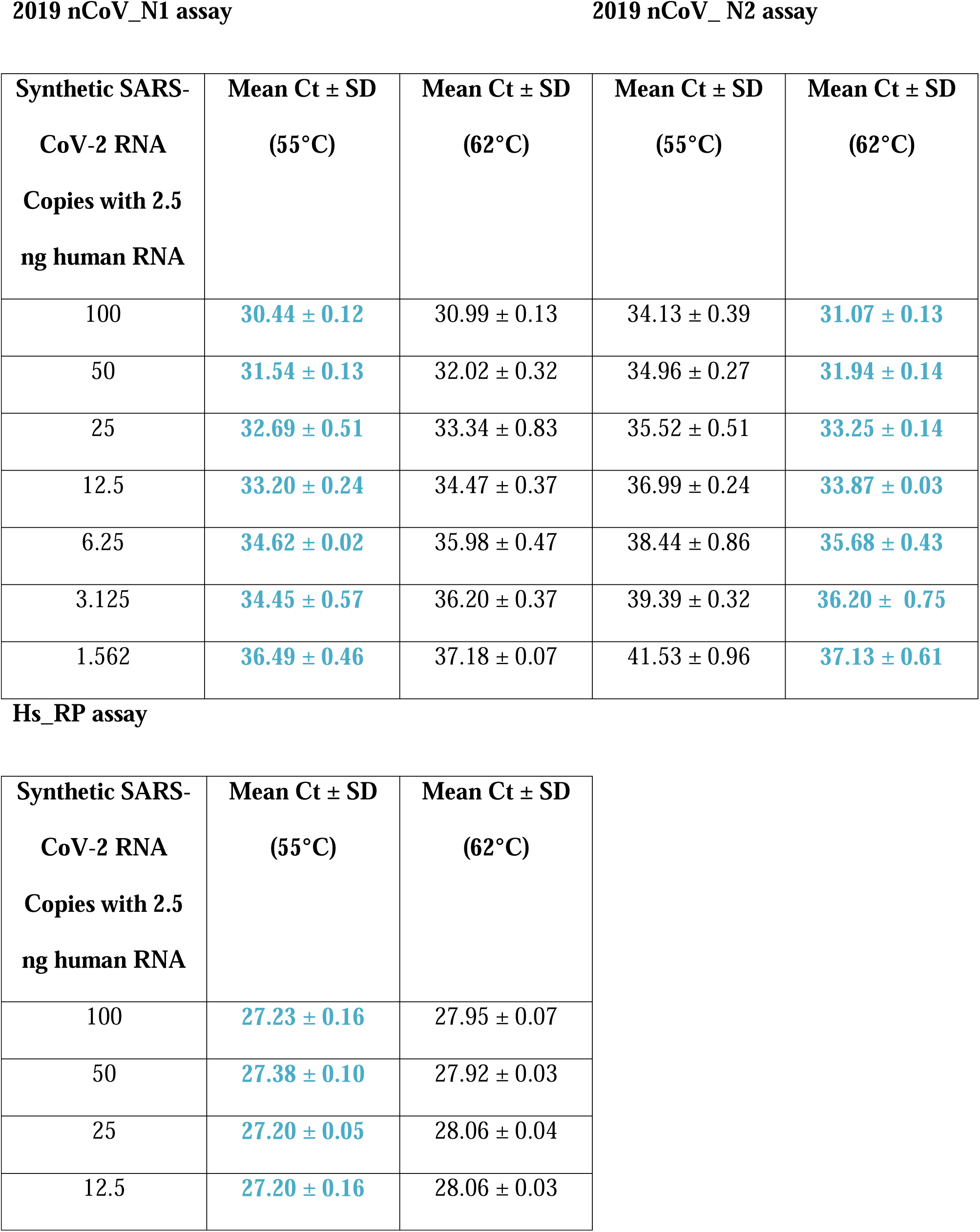
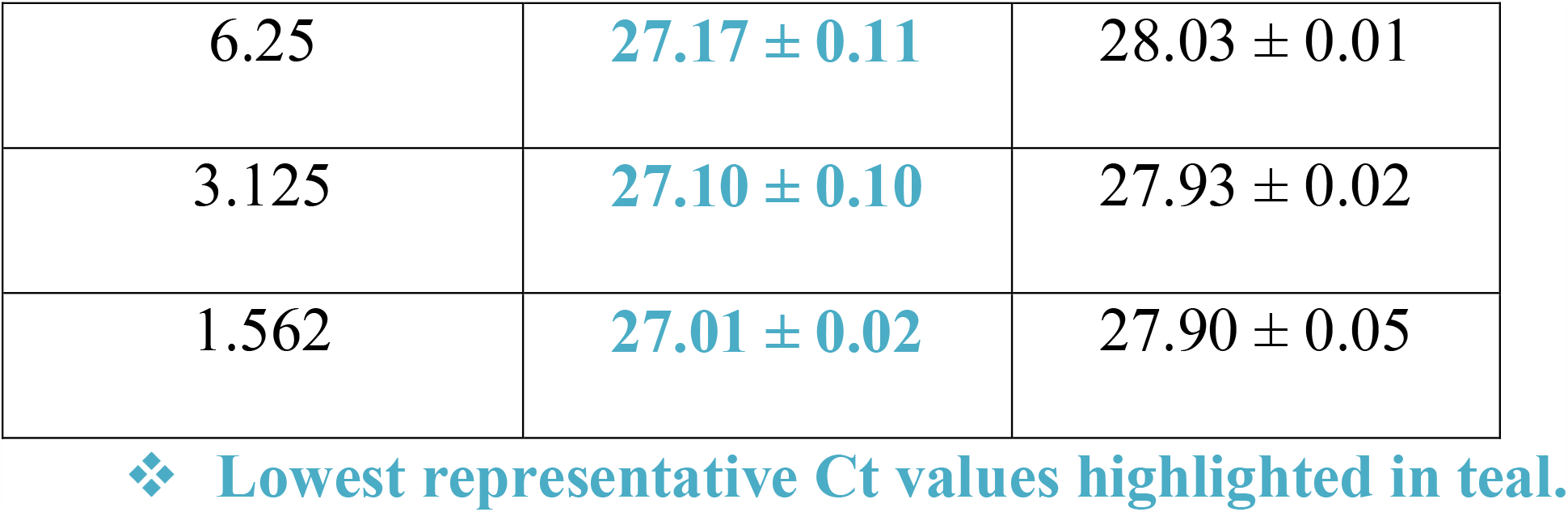
LOD determination without the RNA extraction step.

To evaluate possible effects of viral RNA extraction on the LOD and sensitivity of the assay, we used both manual and automated RNA extractions. In lieu of patient-derived samples, we used a complex matrix of human A549 cell lysate in VTM (Methods section) combined with SARS-CoV-2 synthetic RNA. This matrix was spiked-in with a range of concentrations (103.40, 34.48, 10.34, 3.44, 1.72, 0.86 and 0.34 RNA copies/µl) of the quantitative synthetic SARS-CoV-2 RNA to determine LOD. We did not observe any significant differences in Ct values between manual and automated extractions (Supplemental Table 3). The automated extraction method seemed to be more sensitive for the N1 assay than the manual one, as 0.34 RNA copies/µl failed to be detected when manually extracted. There was no evidence of carry-over between samples during automated extraction, as evident from the negative extraction controls which were VTM alone interspersed between samples during each cycle of QIAcube RNA extractions (data not shown).

Using the lab optimized parameters, we determined the LOD of the COVID-19 diagnostic assay by using a range of quantitative synthetic SARS-CoV-2 RNA serial dilutions spiked into the artificial clinical matrix as described in the Methods section. PCR was done on three biological replicates. As evident from Table 5A, the LOD was found to be 0.86 RNA copies/µl for both the N1 and N2 assays. The intra-assay variance was very low for all the genes: % variance for N1 assay was <1% and for N2 assay was <0.32% (data not shown). The LOD range was then limited to 1.72/0.86/0.34 RNA copies/µl and retested six more times (Table 5B). The inter-assay variability was well within the statistically acceptable range for all the assays: for the N1 assay the variance was <1 % and for the N2 assay variance was <0.30% (data not shown). Finally, the pre-determined LOD of 0.86 RNA copies/µl for this assay was confirmed using 20 biological replicates, as per CDC guidelines (20 separate extractions were done on the QIAcube) [5]. We could detect N1 and N2 assays in all the biological replicates (Table 5C).

**Table 5:** LOD determination with RNA extraction step by QIAamp viral RNA mini kit (Synthetic SARS-CoV-2 RNA spiked into clinical matrix)

**A: Initial LOD range determination**
| **Synthetic RNA**<br>**copies/μl** | **N1_Mean Ct ± SD (No.**<br>**of positives)** | **N2_Mean Ct ± SD (No.**<br>**of positives)** | **Hs_RP_Mean Ct ± SD**<br>**(No. of positives)** |
| --- | --- | --- | --- |
| 103.44 | 30.91 ± 0.17 (3/3) | 30.62 ± 0.16 (3/3) | 30.51 ± 0.11 (3/3) |
| 34.48 | 32.35 ± 0.22 (3/3) | 32.11 ± 0.13 (3/3) | 30.44 ± 0.12 (3/3) |
| 10.34 | 33.99 ± 0.62 (3/3) | 33.17 ± 0.25 (3/3) | 30.55 ± 0.07 (3/3) |
| 3.44 | 36.15 ± 0.92 (3/3) | 35.46 ± 0.48 (3/3) | 30.45 ± 0.09 (3/3) |
| 1.72 | 36.47 ± 1.03 (3/3) | 36.15 ± 0.62 (3/3) | 30.43 ± 0.06 (3/3) |
| 0.86 | 36.77 ± 0.98 (3/3) | 37.01 ± 0.44 (3/3) | 30.30 ± 0.07 (3/3) |
| 0.34 | 37.74 ± 1.11 (2/3) | 37.97 ± 0.47 (2/3) | 30.24 ± 0.11 (3/3) |

**B: LOD determination with limited range**
| **Synthetic**<br>**RNA**<br>**copies/μl** | **Mean Ct ±**<br>**SD (Set 1)** | **Mean Ct ±**<br>**SD (Set 2)** | **Mean Ct ±**<br>**SD (Set 3)** | **Mean Ct ±**<br>**SD (Set 4)** | **Mean Ct ±**<br>**SD (Set 5)** | **Mean Ct ±**<br>**SD (Set 6)** |
| --- | --- | --- | --- | --- | --- | --- |
| **2019 nCoV_N1 assay** |  |  |  |  |  |  |
| 1.72 | 37.81 ± 0.29 | 35.79 ± 0.27 | 35.81 ± 0.77 | 35.95 ± 0.77 | 35.85 ± 0.14 | 34.39 ± 0.13 |
| 0.86 | 37.16 ± 0.34 | 36.39 ± 0.69 | 36.74 ± 0.72 | 36.94 ± 0.64 | 35.04 ± 0.59 | 35.62 ± 0.36 |
| 0.34 | 38.82 ± 0.66 | 36.89 ± 0.57 | 37.53 ± 0.78 | - | - | 36.37 ± 0.89 |
| **2019 nCoV_N2 assay** |  |  |  |  |  |  |
| 1.72 | 36.74 ± 0.21 | 36.13 ± 0.81 | 35.59 ± 0.60 | 36.06 ± 0.55 | 35.34 ± 0.29 | 36.14 ± 0.56 |
| 0.86 | 37.01 ± 0.15 | 36.56 ± 0.21 | 37.58 ± 0.34 | 37.32 ± 0.76 | 36.55 ± 0.69 | 36.88 ± 0.24 |
| 0.34 | 37.97 ± 0.03 | 37.76 ± 0.02 | 37.90 ± 0.45 | 37.74 ± 0.01 | - | 37.42 ± 0.65 |
| **Hs_RP assay** |  |  |  |  |  |  |
| 1.72 | 31.15 ± 0.11 | 31.45 ± 0.02 | 31.30 ± 0.07 | 31.42 ± 0.33 | 31.61 ± 0.32 | 31.10 ± 0.03 |
| 0.86 | 31.23 ± 0.04 | 31.42 ± 0.02 | 31.41 ± 0.22 | 31.47 ± 0.21 | 31.08 ± 0.01 | 31.37 ± 0.09 |
| 0.34 | 31.39 ± 0.05 | 31.29 ± 0.11 | 31.41 ± 0.03 | 30.74 ± 0.07 | 31.04 ± 0.10 | 31.37 ± 0.10 |

**C. Twenty biological replicates of the LOD (0.86 RNA copies/μl in clinical matrix)**
| **N1 assay (Mean Ct ± SD)** | **N2 assay (Mean Ct ± SD)** | **Hs_RP assay (Mean Ct ± SD)** |
| --- | --- | --- |
| 37.16 ± 0.34 | 37.01 ± 0.15 | 31.23 ± 0.04 |
| 36.39 ± 0.69 | 36.56 ± 0.21 | 31.42 ± 0.02 |
| 36.74 ± 0.72 | 37.58 ± 0.34 | 31.41 ± 0.22 |
| 36.94 ± 0.64 | 37.32 ± 0.76 | 31.47 ± 0.21 |
| 35.04 ± 0.59 | 36.55 ± 0.69 | 31.08 ± 0.01 |
| 35.62 ± 0.36 | 36.88 ± 0.24 | 31.37 ± 0.09 |
| 37.16 ± 0.60 | 37.01 ± 0.07 | 30.19 ± 0.04 |
| 36.39 ± 0.02 | 36.56 ± 0.02 | 30.35 ± 0.12 |
| 36.74 ± 0.03 | 37.58 ± 0.39 | 30.37 ± 0.07 |
| 36.60 ± 0.04 | 36.76 ± 0.60 | 31.68 ± 0.05 |
| 38.15 ± 0.35 | 35.64 ± 0.46 | 31.60 ± 0.04 |
| 37.05 ± 0.21 | 36.81 ± 0.22 | 30.88 ± 0.03 |
| 36.32 ± 0.29 | 37.83 ± 0.32 | 31.09 ± 0.10 |
| 36.86 ± 0.25 | 36.81 ± 0.32 | 31.36 ± 0.02 |
| 37.08 ± 0.54 | 36.27 ± 0.72 | 31.57 ± 0.01 |
| $38.44 \pm 0.52$ | $36.74 \pm 0.62$ | $31.15 \pm 0.14$ |
| $36.67 \pm 0.27$ | $37.80 \pm 0.49$ | $31.27 \pm 0.01$ |
| $38.12 \pm 0.31$ | $36.98 \pm 0.21$ | $31.26 \pm 0.04$ |
| $36.19 \pm 0.10$ | $35.97 \pm 0.70$ | $31.45 \pm 0.14$ |
| $36.60 \pm 0.19$ | $36.93 \pm 0.72$ | $31.35 \pm 0.02$ |

## Discussion

In this study we have modified the CDC real-time PCR based assay to detect SARS-CoV-2 to make it more sensitive with the aim to reduce the number of inconclusive results. Modification of PCR parameters decreased the LOD of the assay, which is beneficial for detecting low viral load patient samples. The availability of the complete sequence of the ∼30 kb single-stranded RNA genome of the SARS-CoV-2 virus encoding ∼9860 amino acids early in the pandemic enabled public health organizations and investigating groups to develop highly specific PCR-based diagnostic tests [14, 15]. Investigators have developed tests utilizing varied techniques including multiplexed real-time PCR [16], real-time loop-mediated isothermal amplification [17], and CRISPR-Cas12-based detection [18]. The initial study on the first twelve patients diagnosed with COVID-19 in the US revealed that all patients had detectable levels of SARS-CoV-2 RNA for 2-3 weeks after the onset of illness and that the highest viral load in the upper respiratory tract was present within the first week of illness [19]. This data points toward the necessity for developing very sensitive and specific PCR based diagnostic tests to detect COVID-19 in the acute stages to help in disease management [9, 20].

A number of reports have identified factors that adversely affect PCR results like varying methods of collection of specimens, storage methods, transport medium, shipping time and thermal inactivation to inactivate the virus [21, 22, 23, 24]. A case report of a COVID-19 patient in Wuhan, China, who had four sequential negative real-time PCR test results of his pharyngeal swab before eventually testing positive in the fifth round, points toward the possibility of several factors affecting PCR results negatively [25]. It is quite possible that cases of the recurrence of the disease and initially negative patients turning positive later on, is due to false negative PCR results. False negative results or limitations of PCR-based diagnostic tests can lead to positive COVID-19 patients being diagnosed otherwise, discharged or allowed to leave quarantine and move freely in the society, aiding in transmission of the disease. Even the most widely used point-of-care test in the US, the Abbott ID Now (based on real-time PCR), has been reported to only have an accuracy of ∼85% which led to FDA-issued alerts regarding false negatives [26]. Thus, it is a necessity to derive rapid tests that are not only specific, but also highly sensitive.

In this study we modified the CDC real-time PCR diagnostic assay for COVID-19 in an effort to improve the performance with regard to sensitivity for viral detection and to allow our direct implementation of this tool in a community-based hospital setting. In addition to the list of pathogenic agents that the CDC tested for specificity, we extended the specificity testing to include an additional 27 clinically relevant human pathogenic microbial agents (Supplemental Table 1) and identified no cross-reactivity with any. Modification of the PCR annealing temperature facilitated the development of a more robust assay by lowering the Ct values of the N2 assay in particular (Table 3). An increase in the annealing temperature to 62°C from the CDC recommended 55°C, allowed the N1 and N2 assays to yield similar Ct values that were much lower than the cut-off value of 40 that the CDC recommends as being read as a positive result [5]. The LOD of the modified PCR assay we developed (with the inclusion of the extraction step) is 0.86 RNA copies per µl (Table 5), which is more sensitive than the CDC assay. According to the CDC EUA protocol, the LODs of the N1 and N2 assays (with the Qiagen EZ1 DSP extraction kit) are 1 and 3.16 RNA copies/µl, respectively [5]. Without the viral RNA extraction step, the LOD was found to be 1.562 copies of the SARS-CoV-2 synthetic RNA for both N1 and N2 assays under the lab optimized conditions (Table 4). However, under the CDC recommended PCR conditions, the N2 assay had a LOD of 3.125 copies of synthetic RNA (Table 4).

The SARS-CoV-2 virus has been detected in several types of biological specimens including sputum, saliva, feces and semen [27, 28]. A recent study has reported sputum to be the most accurate sample type for COVID-19 diagnosis, followed by nasal swabs and bronchoalveolar lavage fluid [29]. To our advantage, PCR based tests can be adapted to suit any type of starting material as long as good quality viral RNA can be extracted. The CDC recommends that a PCR Ct value of <40 is to be considered as positive when interpreting diagnostic results and if either the N1 or the N2 assay is positive, the result should be interpreted as inconclusive [5]. By modifying the assay to get lower Ct values of the N1 and N2 assays even for low viral load samples, we have tried to reduce the number of inconclusive results. This modification will have a tremendous impact when applied in a clinical setting to manage a pandemic of this magnitude.

## Data Availability

The data is available immediately upon publication.

## Acknowledgments

The authors would like to acknowledge the technical advice and suggestions received from Dr. John A. Martignetti and Dr. Olga Camacho-Vanegas of Icahn School of Medicine at Mount Sinai, New York. The authors are also thankful to Ms. Sucheta Godbole of Rudy L. Ruggles Biomedical Research Institute, Nuvance Health for help in statistical calculations.

## Figure Legends

**Supplemental Figure 1. Dynamic linear real-time PCR range with CDC-recommended conditions**.

The linearity range of 2019 nCoV_N1, 2019 nCoV_N2 and Hs_RP Taqman assays was determined with CDC-recommended PCR parameters and various concentrations of positive plasmid control DNA by real-time PCR.

